# GENDERED GAPS TO TB PREVENTION AND CARE IN KENYA: A POLITICAL ECONOMY ANALYSIS STUDY

**DOI:** 10.1101/2023.07.20.23292959

**Authors:** Leila Abdullahi, Sandra Oketch, Henry Komen, Irene Mbithi, Kerry Millington, Stephen Mulupi, Jeremiah Chakaya, Eliya Zulu

## Abstract

**Background:** Tuberculosis (TB) remains a public health concern despite the massive global efforts towards ending TB. Kenya is among the high burdened countries globally with the TB prevention and care efforts hampered by poor health systems, resource limitations and other socio-political contexts that inform policy and implementation. Notably, TB cases are much higher in men than women, and therefore, the political economy analysis study provides in-depth contexts and understanding of the gender gaps to access and successful treatment for TB infection.

**Methods:** The Political Economy Analysis (PEA) adopted a qualitative in-depth approach that targeted 16 Key Informant Interviews (KIIs). The Key Informant Interviews (KIIs) were distributed among government entities, academia, non-state actors, and community TB groups.

**Results:** The themes identified were mapped onto the applied PEA analysis framework domains. The contextual and institutional issues included gender concerns related to the disconnect between TB policies and gender inclusion aspects, such as low prioritization for TB programs, limited use of evidence to inform decisions and poor health system structures. The broad barriers influencing the social contexts for TB programs were social stigma and cultural norms such as traditional interventions that negatively impact health-seeking behaviors. The themes around the economic situation were poverty and unemployment, food insecurity, and malnutrition. The political context centered around the systemic and governance gaps in the health system from the national and devolved health functions.

**Conclusion:** Overall, broad contextual factors identified from the political economy analysis widens the disparity in targeted gender efforts toward men. Following the development of effective TB policies and strategies, it is important to have well-planned gendered responsive interventions with a clear implementation plan and monitoring system to enhance access to TB prevention and care.

**Strength and limitations:** The study provides insights into the political, economic, and social contexts influencing TB prevention and care efforts. Additionally, to the best of our knowledge, this will be the first PEA to evaluate the TB program from a gendered perspective.

As a limitation, the study is missing the voices and lived experiences of men leaving with TB as this was not part of the study scope. As such, we do not have their perspectives as they may better articulate some barriers faced with access and uptake of TB prevention and management programs.

## BACKGROUND

Globally, tuberculosis (TB) is among the leading causes of mortality in developing countries with an estimated 10.6 million people that developed TB and 1.6 million deaths in 2021[1]. The TB cases were notably higher in the male compared to the female gender [1, 2] at 56.5% and 32.5%, respectively, while the prevalence for children was 11% [1]. Kenya is ranked 13^th^ among the 22 highly burdened countries [3–5] and is exceedingly burdened by TB, TB-HIV co-infection, and multi-drug resistant TB [1, 4, 5]. In 2021, the estimated TB incidence in Kenya was 133,000 while TB mortality was 31,000 [1] with an overall disproportionately high burden in men comprising 64% of all TB cases [6].

The WHO End TB strategy endorsed in 2014 has set the goal for ending the TB epidemic globally by 2035 with a targeted 95% decline in TB deaths and a 90% reduction in TB incidence [7]. TB prevention and care are hampered by delayed diagnoses and treatment, poor health infrastructures, and under-reporting for TB cases [8]. Consequently, the highly burdened TB countries need to continuously focus on the priorities for TB prevention and care, such as health systems strengthening, optimizing the linkage to treatment through meeting the needs for TB services [9].

TB treatment involves taking anti-tuberculosis drugs for at least six to ten months which includes an intensive phase comprising the first two to three months that has a combination of a two-dose regimen and followed by a continuation phase of four or six months which has a one-dose regimen [10, 11]. However, the treatment regimen for treatment failure is up to 18 months, including the first six months of the intensive phase and 12 months of the continuation phase [10]. TB patients who are undiagnosed or have poor adherence to treatment are highly infectious. Moreover, the patients with poor adherence are likely to either relapse or succumb to TB [12].

There have been efforts to increase access, acceptability, adherence and uptake for TB prevention and care programs to the general population and among children, including improved diagnostics [13, 14], capacity building of health care providers [15] use of mobile and digital health technologies [16], integrated screening for TB and COVID 19, TB self-testing, active case finding and contact tracing, networks for TB survivors and TB communities [10, 17–20]. Although there have been global and country-level efforts to control TB infections, the ultimate success depends on concerted efforts around targeted populations, including a gendered approach while designing TB prevention interventions [21, 22]. Despite the effort to have TB gender-disaggregated data, there is a paucity of information on gender-responsive interventions to address the significantly high cases of TB among men.

Through the Leaving no-one behInd; Transforming Gendered Pathways to Health for TB (LIGHT) Consortium ongoing work in Kenya, the African Institute for Development Policy (AFIDEP) and Respiratory Society of Kenya (ReSok) conducted a PEA study to understand the barriers to access and successful treatment for TB infection among Kenyan male adolescents and adults. The PEA study seeks to dig deeper into the gaps that lead to TB treatment interruptions and poor health-seeking behaviors among men and recommends approaches to enhance TB gender-responsive programs to improve TB access and successful treatment among men.

## OBJECTIVE

To understand the political and economic contextual factors that hinders access to and successful treatment for TB infection among Kenyan male adolescents and adults.

Specific objectives

i. To understand the contextual factors that hinder access to and successful treatment for TB infection among Kenyan male adolescents and adults.
ii. To explore the political and power dynamics that shape opportunities and challenges towards TB gender-responsive programs.

## METHODOLOGY

### Study design

The PEA methodology is a structured qualitative approach examining power dynamics and economic and social forces influencing development. The analytical approach is informed by applied political economy

**Figure 1:**
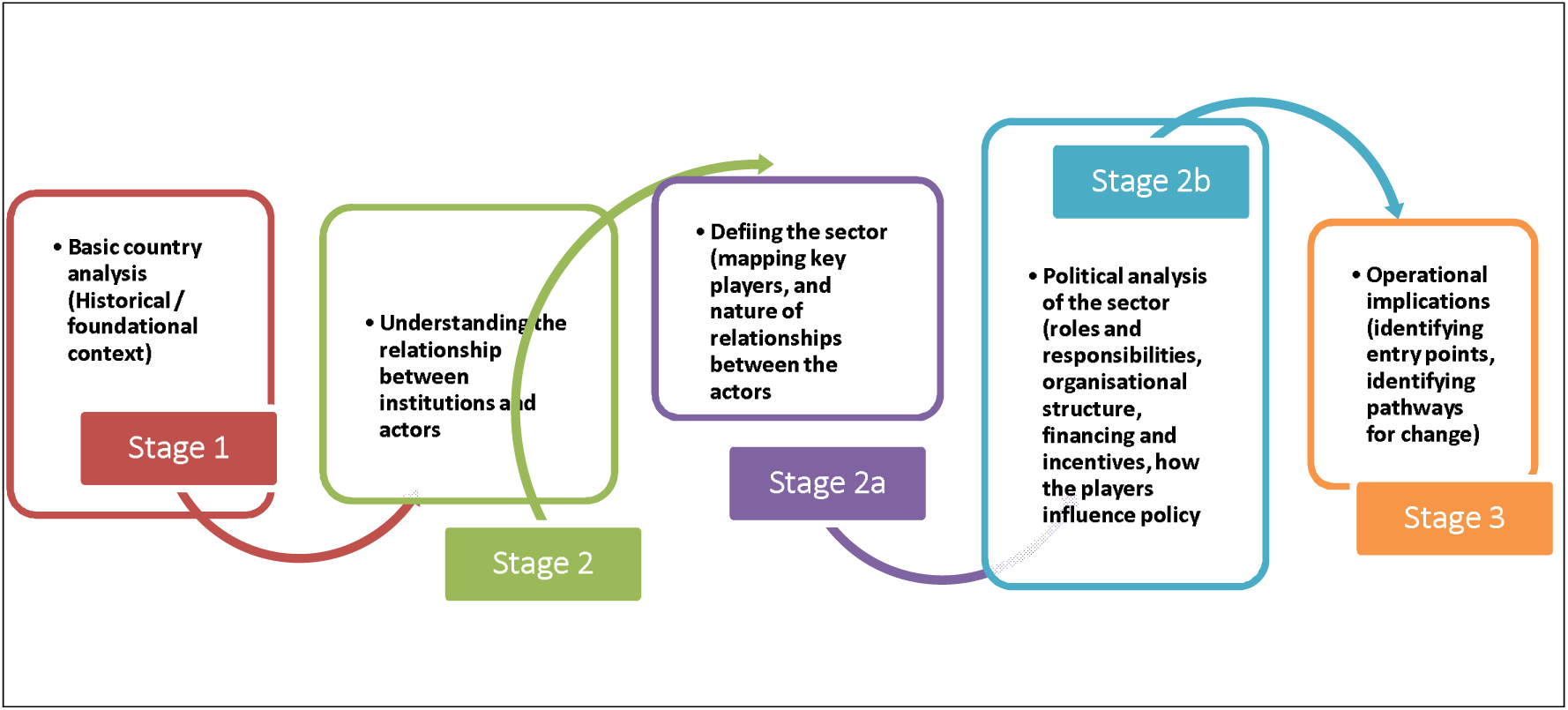
Conceptual framework for the PEA. Source: Adapted from Applied political economy analysis framework [25]

### Study site and population

Through consultations with the National TB programme implementation team, a list of study participants was generated as a starting point for data collection comprising national and county level top- and mid-level health policymakers in the TB ecosystem. The target sample included TB actors in Kenya who works alongside and/ or influence the policymakers on approaches to improve access to and successful treatment of tuberculosis infection and incidence among Kenyan male adolescents and young adults.

### Sampling

In Jan 2023, 16 participants were purposively sampled all over Kenya using purposive, maximum variation sampling technique. The participants included health-sector-related government entities, civil society groups in health, individuals in research institutions conducting health-related research, the private sector, academia with expertise on Kenya’s political economy, TB coordinators, and, TB champions. Below is the summary of the stakeholders interviewed and their position (**Table 1**).

**Table 1:**
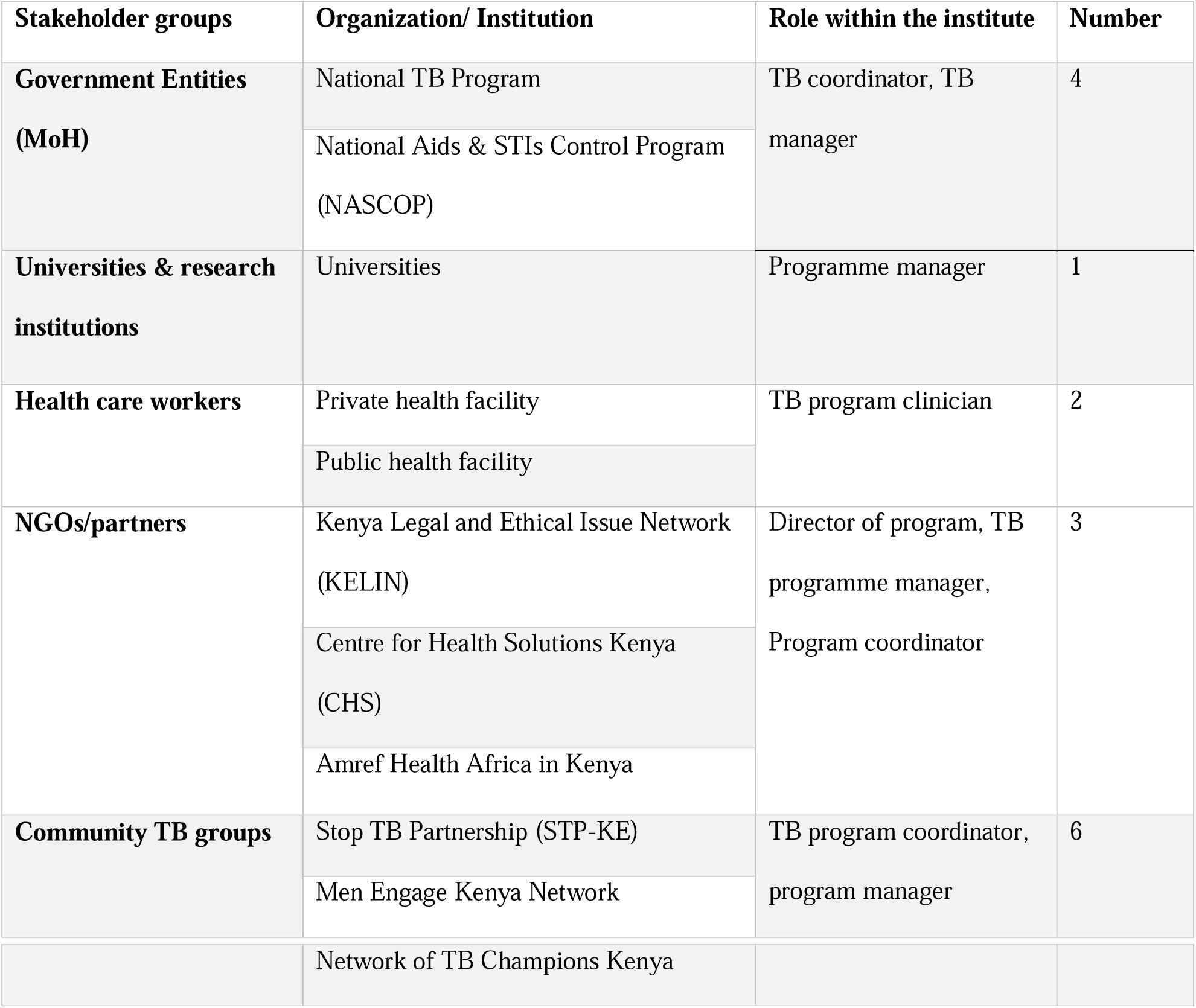
Stakeholders Interviewed.

### Data collection

Data were gathered from two main sources including: desk review of LIGHT programme documents, MoH and research institutions and an in-depth interview to include PEA lenses. The desk review helped shed light on the dynamic interaction between power, institutions, and actors and how these facilitate or constrain interventions aimed at reducing the number of people affected by tuberculosis (TB) in the country.

### In-depth interviews

A semi-structured interview guide was designed guided by the PEA framework [25] to capture information related to interaction (between power, institutions, and actors) between political interests and power dynamics that shape interventions around gendered TB and opportunities and pathways for greater impact in reducing the number of people affected by tuberculosis (TB) in the country. The semi-structured interview guide had a mix of open- and closed-ended questions. Interview participants were recruited and followed up using email and telephone. All the interviews were conducted virtually using Microsoft Teams. All the interviews were audio recorded, and transcribed after obtaining written informed consent. The interview transcripts were anonymised and stored in a password-protected file accessible only to the project team.

### Data analysis

Data from the qualitative interviews were analyzed using a combination of inductive and deductive approaches utilizing the framework thematic approach [23, 24]. Transcripts were coded using the Dedoose software by two members for joint review through a series of meetings. The codes were then mapped onto the domains from the applied PEA analysis framework [25] while identifying emerging themes and sub-themes.

## RESULTS

### Contextual and Structural issues

#### Gender concerns related to TB care

Men account for roughly twice the TB burden while displaying extremely low health-seeking behaviors with the highest TB treatment interruptions [26–28]. Further, men with a TB infection face unfavorable outcomes of TB deaths and treatment failure possibly due to non-adherence to treatment [26, 28]. The qualitative inquiry reported on the concerns around limited targeted interventions towards men which in turn do not meet the needs of men, thereby influencing their overall access to TB services.

> *“Most of the time, more men than women get tuberculosis, which is a problem. …. I agree that most women now are more empowered but they’re leaving out men”.* (KII, TB Champion, 2022)

> *“…, there are minimal interventions with TB in Kenya geared towards men. We don’t focus on men regarding case finding, meeting their needs, and so on, which is quite unfortunate”.* (KII, Development Partner, 2022)

Kimani et al 2021 recommended that actors should actively target men with systematic TB screening and diagnostic services to ensure early diagnosis and interrupt transmission [29]. The recommendation aligns with the majority of the key informant responses where participants emphasized the need to invest in male-friendly diagnostic and screening services, with the aim of reducing undiagnosed TB.

> *“The social workplace and networks of TB stakeholders are increasingly being taken into consideration rather than visiting men at their homes”.* (KII, TB Coordinator, 2022)

> *“Since men are more likely to be affected than women, we usually focus on places where men gather, such as social places”.* (KII, Development Partner, 2022)

> *“Focusing on men gathering places as potential intervention spaces is important if we are to reduce the burden of TB and achieve the End TB targets”.* (KII, MoH,2022)

#### Health infrastructure

In Kenya, there exist broad TB diagnostic tests in urban compared to rural settings thereby higher costs in the health facilities in urban areas [27, 30, 31]. Further, the public health care system is overworked and with very high waiting times thereby limiting access to the impoverished population that would otherwise seek care in these facilities [32]. Additionally, there are reported logistical issues around access to TB services where the health facilities are in far-off areas [31]. Therefore, a need to decentralize treatments offering TB services [33]. Overall, the high TB treatment costs is attributed to a wide range of factors such as laboratory costs, drug prices, nutritional support and hospitalization costs [30]. The participants from the qualitative finding echoed the need to have TB services more accessible by equipping more hospitals with the necessary resources and offering flexible times to meet the needs of patients beyond the 8 am-5 pm working hours. These efforts would largely lead to the expansion of TB services to men who are casual laborers who work during the day.

> *“Some facilities do not offer TB services as expected, there is a need to have more healthcare staff and equipped facilities in Kenya offering TB services”.* (KII, Health care practitioner, 2022)

> *“The Arid and Semi-Arid Land area harbors vulnerable and pastoralist communities that have additional challenges to access health care due to the fixed hospital working hours of between 8am to 5 pm. This timing limits casual laborers who are faced with the competing needs of work to provide for their families against seeking medication”. (*KII, TB Champion, 2022)

> *“Participants recommended that health centers offering TB services be flexible and offer services on the weekend to suit everyone”.* (KII, policy maker,2022)

### Social and Economic Context

#### Social and cultural norms

In Kenya, the high TB burden mostly in men [26] is partly attributed to poor health-seeking behavior which is attributed to cultural norms and traditional beliefs. The cultural beliefs lead to seeking traditional interventions including a preference for alternative medicine thereby taking longer to get medical help or TB treatment attrition [26, 31, 34–36]. Furthermore, social stigma attributed to TB and HIV/AIDS increases patients’ delay to treatment [34].

> *“There is a stigma attached to TB and is scary. …If you delay TB diagnosis, you become slim and thin and people mistake it for HIV. The stigma associated with HIV and TB has created late case detection or missed cases”. (*KII, TB champion, 2022).

#### Poverty and Unemployment

The Africa Development Bank (AfDB) reports that the inflation in Kenya was at 6.1% in 2021 with a public debt surge of 68%. Additionally, the proportion of Kenyans living in extreme poverty was 16%, and unemployment at 12.3% [37]. Consequently, poverty makes it easier for TB to spread, mainly because it affects living conditions (people living in overcrowded, poorly ventilated homes) [38]. Further, poverty leads many people to hesitate medical treatment and seek cheap alternate treatments such as herbal medications [39].

In Kenya, unemployment is a major contributor to TB treatment interruptions and lost to follow-up [26, 27]. Consequently, men who are the sole breadwinner and rely on daily wages face the hurdle of providing for the family or seek care for ailments or worrying symptoms [26]. The study participants emphasized on the unemployment effects on to poor health outcomes such as the inability to pay for medical care while ill and the usage of drugs at reduced dosages necessitated by the logistical and access costs, which leads to drug resistance in the body. Worthwhile to note, even though TB treatment is free in public health facility, however, the 8-5 pm working hours hinder the men from seeking care as they are working to obtain their daily wages.

> *“Unemployment affects health because if you have no money, you cannot go to the hospital when sick. Also, you can’t afford drugs you go for half doses creating drug resistance in the body”.* (KII, Development Partner, 2022)

Further, the majority of the participants described the poor state of the economy which has been exacerbated by the COVID-19 pandemic which led to the loss of jobs and high costs of living.

> *Most people lost their jobs after corona and now jobs are very scarce impacting TB access.* (KII, Health care practitioner, 2022)

#### Food insecurity and malnutrition

Like other countries, Kenya is affected by climate change brought on by global warming thereby affecting agriculture and food security [40, 41]. Food has a long-term impact on health since it can lead to malnutrition and mental health [40, 41]. TB patients with severe malnutrition have higher risks of failure rates or deaths [28] and it is a key driver of the TB epidemic in Kenya [42]. Therefore, a substantial contributor to TB in Kenya is food insecurity. The interviewed participants agreed that most Kenyans, including males, focus on food and daily sustenance rather than healthcare since it has reached a point where it is equated to a life-or-death situation.

> *“…., because of global warming, we are experiencing a lack of rain, leading to drought and famine, so everyone is more concerned with food and daily nourishment than health care”.* (KII, Development Partner, 2022)

### Political Context

#### The country and its devolved counties

Since 2010, due to the devolved system, each county in Kenya has had a different level of efficiency in providing healthcare services [43]. The qualitative inquiry findings showed that the counties are struggling and need to improve the situation regarding their decisions, which is especially low in TB programs. Most counties indicated that the national government only provides them with meager funding, with a further low prioritization on TB programs by the county health teams. Additionally, most participants indicated that corruption and lack of coordination between national and county government plays a major role towards poor health service delivery.

> *“The health sector is really struggling because of how the health services are being run. The counties are having a hard time, and the national decision-making process isn’t making things better at the county level”.* (KII, Policymaker, 2022)

> “*They get a lot of money and allowances, which they use to benchmark in places like Dubai. And sometimes they choose to buy furniture …”.* (KII, Health care practitioner, 2022)

> *“…. They must depend on the national government. We are behind other countries in finding cases because of this. Based on the report from last year, we didn’t diagnose close to 50% of the people we were supposed to”.* (KII, TB Coordinator, 2022)

### Institutional environment

#### TB policies and gender

Overall, TB policies and strategy implementation in Kenya have been hampered by limited budgetary allocations [44]. The policy analysis revealed that various components of active TB including management and diagnosis were included in Kenya’s National Strategic Plan for TB and Lung Health 2019-2023 [45]. However, the policies/strategies lack a gendered approach to address men’s barriers to TB care. These findings were validated by the qualitative study which pointed out the overlook from policymakers and implementers on the gender influences on tuberculosis and its control.

> “*We have policies and guidelines, but none talk about a specific gender. Instead, they cover everyone. But we’ve found that TB affects men more than women. This is a hot topic, and we need to consider gender issues in TB care and prevention*.” (KII, Policymaker, 2022)

It is evident from the data that men are more burdened with regards to the highest TB mortality

[1, 2] with minimal interventions targeting men. The consideration for a gendered interventions/approach should stem from the National TB programme and partners meetings, where there is strengthened advocacy for targeted gendered approaches for TB care and prevention among young males and elderly persons.

> “*Based on survey* results*, men are more affected and wait longer to get help. So, TB is different for men and women in some ways. So, TB policy planning and advocacy should include both men and women, …, you should focus on other ways to reach them*.” (KII, Academician, 2022)

#### Evidence-Informed Decision-Making capacity gap

Skills in evidence synthesis, utilization, and communication are vital among policymakers, Civil Society Organisations (CSO), researchers, and media to communicate findings to stakeholders including the community [43]. Most of the research participants identified capacity gaps in the translation of evidence that hinders the dissemination of research findings to inform healthcare decision-making. Respondents from the development partner suggested an online TB research hub to disseminate research findings in a plain language summary to aid decision-makers in their decision-making process.

> *“…, but most of it [research] doesn’t get to the people who make policy because it isn’t shared with them. They can’t use it to decide what to do or develop policies. So, if there were, you know, research hubs, I think that would go a long way toward helping us get all this information together*”. (KII, Development Partner, 2022)

Additionally, participants concurred to the need for the researchers to break down technical research information for easy consumption by all actors who use evidence for decision making such as policymakers, and legislators, among other key stakeholders.

> “…*investing in the capacity on communication strategy will help to deliver the dedicated TB messages to policymakers and the community at large”*. (KII, Development Partner, 2022)

Importantly, the research participants indicated the need for a dedicated staff member within the government sector who will support the coordination and collaboration of the government TB sector with research institutes including universities, development partners, and NGOs, where a number of innovative research are ongoing. Through increasing capacity, young upcoming researchers can receive mentorship from senior researchers on emerging technologies for TB management and diagnosis. Importantly, through knowledge-sharing platforms like digital technology, the community can gain awareness and knowledge while dispelling pre-existing myths and prejudices concerning TB.

> “*The problem is to do with resources. There is a need to add more resources to the research sector. They need to have specific staff that deals with research. Collaboration between organizations, e.g., universities, will also work*”. (KII, Healthcare Practitioner, 2022)

### TB Actors in Kenya

TB as a public issue in Kenya that has attracted a diverse set of actors in government (at the three levels), non-state actors, and development partners. The degree of involvement of these actors is informed by their interests, roles and power relations in the TB control program. Overall, the Ministry of Health plays a central role in the management of TB in the country. The National Tuberculosis and Leprosy Control Programme (NTP) controls most of the funding for TB work in the country. The National level actors including the researchers, community, healthcare professionals, TB champions, development partners, civil society actors, and politicians are occasionally engaged in various forums, including TB Technical Working Group (TWG), on the issue TB care and prevention. However, an action plan with gender-responsive approaches is still missing.

#### Roles and power relations

The Directorate of Preventive and Promotive Health in the Ministry of Health provides the overall leadership on TB prevention and care [46]. The NTP provides a key interface between policy, strategic planning, and practice, where they offer technical assistance and decisions around TB prevention and care at the national and county level [47, 48]. Furthermore, The NTP has had significant reforms in the past decades where activities aimed at ending TB have been decentralized to the county level (previously known as the district level). Consequently, the decentralization process has strengthened the county-level power and responsibility for priority setting, budget allocation, and training [47, 48]. To note, the resource and allocation of health resources is both a technical and political process [49]. The county health management teams spearheaded by the county TB coordinators develop the TB budgets and work plans while the political leaders are included in the resource allocation processes [49]. However, there exists political interference around the health sector utility and budgeting at the county levels [47].

Kenya is heavily reliant on Development partners for TB financing [48] with reported challenges on limited funding and poor collaborations of TB programs with the private health sectors [50, 51]. Development partners have largely contributed to the annual global funding towards ending TB with reported funding gaps in recent years [48, 52] and have pushed for adoption of program-based budgeting to facilitate resource allocations and prioritizations [53, 54].

The qualitative findings show an urgent need for the government to prioritize TB funding in the country. “*Up to 80% of TB funding is from donors, thus risking sustainability”*.

> *“If NTP doesn’t get enough money from the national government, which is especially true now. With some donors giving less money, it’s easy to see how this could be a big problem for the country”.* (KII, TB Coordinator, 2022)

In Kenya, few NGOs like The Kenya Legal & Ethical Issues Network on HIV and AIDS (KELIN) have segregated gendered guidelines and services where they offered gendered services in a structured way. KELIN organization concentrate on specific interventions involving men in the workforce. The organization has an initiative that goes to areas where men are available, including their leadership. Some of the interventions include performing screenings and health education.

> “*Our guidelines call for different kinds of service delivery. Since men are more likely to be affected than women, we usually focus on places where men gather, such as social places*”. (KII, Development Partner, 2022)

> “*Stigmatized men are being attended to during specific times of their convenience. Because health-seeking behavior among men is poor, they are being attended to; awareness, screening, and delivery of medicines at their convenient places”.* (KII, TB Champion, 2022).

> “*Men should be targeted where they are, rather than waiting for them to come to treatment centers, to ensure they do not miss out on their jobs while seeking or receiving care. They do not forfeit their pay*”. (KII, TB Coordinator, 2022)

The Stop TB Partnership and TB champions have played a significant role in raising awareness regarding gender and human rights through their Communities, Rights, and Gender (CRG) initiative. The interventions help to find cases, raise awareness, or participate in any community-based TB response. The organizations actively engage. with funders, including the Global Fund and the US government, among others, to mobilize financial resources for TB initiatives and to undertake community awareness and dialogues.

> “*We have been making the people we work with more aware. We look at it regarding gender and human rights. We have taught them about gender and human rights so that when they go out into the community to look for TB cases, raise awareness, or make any other community-based TB response, they are aware of the gender and rights of the community”.* (KII, TB Coordinator, 2022)

Communities have the potential to improve overall TB treatment outcomes at minimal costs [55]. Additionally, the inclusion of TB care and other health care in the communities largely improves acceptance and access [55, 56]. The qualitative inquiry reported that most communities are slowly changing and are becoming positive and receptive to TB programs involving TB policies, including men, women, and the community at large. Further, there’s a need to focus on more sensitization, awareness, and capacity building of community members, Community Health Volunteers (CHVs), TB champions, healthcare workers, and policymakers.

> “*Most communities are changing and are open to TB programs, …… To amplify further, the focus needs to be on raising awareness, lobbying for resources, and leadership, and getting everyone on board*.” (KII, TB Champion, 2022)

Civil society organizations have a keen interest in ending TB and incorporated community interventions including the use of TB champions to create awareness.

> “*The social workplace and networks of TB stakeholders are increasingly being taken into consideration rather than visiting men at their homes. Community Rights and Gender (CRG) is being included as a subject area by the Network of TB Champions as it advances from the entire cascade regarding prevention, care, and cure*”. (KII, TB Coordinator, 2022)

## DISCUSSION

Reducing the global TB burden requires concerted efforts such as a focus on gender, institutional and socio-political lens that are essential aspects in the TB prevention and care cascade that will ultimately contribute to efficient and effective programming. There were notable concerns around gender disparity with high TB treatment interruptions and default among the male compared to their female counterpart [57, 58]. This study further noted the need for targeted gender-responsive interventions and development of progressive policies to improve men’s health seeking behaviors. Additionally, the targeted interventions need to consider structural system such as surveillance of loss to follow-up, decentralized TB services that actually reach even the marginalized populations, and reduce other opportunity costs associated with the treatment. The need for targeted gender-responsive TB interventions is consistent to other study findings, including a study done across highly burdened TB countries which reported on the need to consider efficient mechanisms in the policy and administration, including innovative financing policy options for successful gender-responsive TB implementation guidelines and policies [59]. Moreover, mitigating the opportunity costs associated with treatment, such as time loss, income, long distance to treatment and household expenditures is a consistent finding from other settings [60] which leads to increased access to health services.

The Rio political Declaration held in 2011, emphasize on the need to address inequalities and inequities around the social determinants of health including giving special attention to gender-related aspects [59]. One key social determinant impeding TB treatment and completion are socio-cultural beliefs, such as the use of alternative medicines and social stigma have been reported in various studies [61–64]. There is an urgent need to necessitate culturally sensitive health education on TB disease targeting men including clarifying any myths and misperceptions and enhancing positive attitudes towards TB disclosure. The second reported determinant that hampers TB treatment and care is poverty which, is concurrent with other reported findings [65, 66]. This therefore stresses on the need for good public health strategies [67] and socio-economic empowerment interventions [66]. Furthermore, the negative impacts from the global warming call for healthcare systems preparedness in dealing with the health impacts of the climate change like malnutrition due to drought. However, the majority of the countries including Kenya are not sufficiently prepared [40]. This, therefore, calls for concerted efforts around the country’s financial, logistical and systemic response.

This PEA found that evidence-informed decision-making is a key strategy to inform decision towards sustainable efforts towards ending TB. Skills in evidence synthesis and utilization are vital among governments, research, and implementing partners to inform the decision-making process [68, 69]. To achieve this, research needs to be prioritized at the country and regional level with adequate dissemination capacity to inform policy and practice [70–72]. These often times are impeded by different political prioritizations such as poor governance, limited financial and budgetary prioritizations [59].

On the other hand, the researchers need to assist TB implementers in identifying the issues contributing to gender disparity in seeking health care. Importantly, Kenya must adapt its resource-allocation culture to ensure the planned gender-responsive strategies are funded to ensure successful implementation and monitoring. The government and other TB stakeholders can brainstorm on interventions to combat TB in a gender-transformative manner. For example, the government can put the already developed policies into practice with the necessary human and financial resources.

## CONCLUSION

This PEA study demonstrates an interplay of politics and other socio-dynamics in the health decision-making and implementation space. Overall, there exist broad contextual factors identified from the political economy analysis that ultimately widens the disparity in targeted gender efforts toward men. Following the development of effective TB policies and strategies, it is important to have well-planned gendered responsive interventions with a clear implementation plan and monitoring system to enhance access to TB prevention and care. Moreover, capacity strengthening among researchers and policymakers on how to translate evidence into an actionable recommendation is vital to inform decision-making.

Recommendations:

- TB stakeholders need to include gender-responsive plans, interventions and strategies in existing policies to address gender disparities in TB care and treatment cascade.
- There is an urgent need to mobilize, and earmark resources to enable the successful implementation and sustainable gender-responsive approaches for TB care and treatment.
- Stakeholder engagement and collaboration are critical to implementing effective gendered TB policies to end TB. Most importantly, engaging with communities can establish a better understanding of real-life issues to inform evidence-based policies that address the specific needs of men and women.

## Data Availability

All data produced in the present study are available upon reasonable request to the authors

## List of abbreviations

AIDS: Acquired Immune Deficiency Syndrome
CHVs: Community Health Volunteers
HIV: Human Immuno-deficiency Virus
KII: Key Informant Interview
KELIN: Kenya Legal & Ethical Issues Network on HIV and AIDS
PEA: Political Economy Analysis
TB: Tuberculosis
WHO: World Health Organization.

## Declarations

### Ethics Approval and consent to participate

The qualitative study obtained ethics approval from Kenya Medical Research Institute (SERU number 4205). All participants provided written informed consent before participation in the study.

### Consent for publication

Not applicable.

### Availability of data and materials

The data used and/or analysed during the current study are available from the corresponding author on request.

### Competing Interests

The authors declare that they have no competing interests.

### Funding

UK aid provided funding for this research. The content is solely the responsibility of the authors and does not necessarily represent the official views of UK aid. The funders had no role in the design of the study and data collection, analysis, and interpretation of data and in writing the manuscript or decision to publish.

### Authors contributions

All study authors were involved in the conceptualization of the study. EZ and JC provided overall guidance on study design and manuscript preparation. LA was involved in project management. IM, HK and LA were involved in data collection, data coding and analytic support, and manuscript preparation. All authors were involved in drafting various sections of the initial manuscript. All authors read and approved the final manuscript.

## Acknowledgements

The study was part of the LIGHT programme. LIGHT is a cross-disciplinary global health research programme funded with UK aid from the UK government in partnership between the UK and 6 partners institute: African Institute for Development Policy (AFIDEP), Kenya and Malawi; Zankli Research Center - Bingham University (ZRC), Nigeria; Malawi-Liverpool-Wellcome Trust Clinical Research Programme (MLW), Malawi; Respiratory Society of Kenya (ReSOK), Kenya; Makerere University Lung Institute (MLI), Uganda and Liverpool School of Tropical Medicine (LSTM) Leaving no-one behind; transforming Gendered pathways to Health for TB (LIGHT) Consortium.

## Notes

### Competing Interest Statement

The authors have declared no competing interest.

